# Association of Diet Quality and Physical Activity through Young Adulthood With Mid-Life Cardiovascular Health Trajectories: The Coronary Artery Risk Development in Young Adults (CARDIA) Study

**DOI:** 10.64898/2026.10.19.26365962

**Authors:** Ishita Rai, Hongyan Ning, Jing Li, Kelley Pettee Gabriel, Donald M. Lloyd-Jones

## Abstract

**Background:** Young adulthood is a critical life course window during which cardiovascular pathophysiology develops, yet diet quality and physical activity (PA) are typically studied as separate, static exposures. We examined joint associations of diet and PA across young adulthood with midlife cardiovascular health (CVH) patterns.

**Methods:** Among 2,806 participants in the biracial Coronary Artery Risk Development in Young Adults (CARDIA) cohort, group-based dual-trajectory modeling identified latent classes of joint Dietary Approaches to Stop Hypertension (DASH) score and PA from exam Year 0 to Year 20 (mean age range at each exam 25–45 years). Group-based trajectory modeling defined midlife CVH trajectories using Life’s Essential 8 (LE8) scores (5 physiologic components) from Year 20 to Year 35 (mean age range at each exam 45–60 years). Multinomial logistic regression, adjusted for age, sex, race, and education, related young-adult behavioral groups to midlife CVH status (high ≥80, moderate 50–79, or low <50).

**Results:** Four joint diet/PA groups from young adulthood behaviors were identified: low DASH/very low PA (Group 1; 10.9%); low DASH/moderate-declining PA (Group 2; 25.6%); moderate DASH/high-declining PA (Group 3; 41.5%); and high DASH/high-declining PA (Group 4; 22.0%). Three non-overlapping midlife CVH trajectories (low, moderate, high) emerged. Compared with Group 1, participants in Groups 2, 3, and 4 had 4.8-, 8.6-, and 21.7-fold higher odds of high versus low CVH. The joint odds ratio (21.7) exceeded those for high diet quality (7.2) or high PA (10.3) individually. Associations persisted after adjustment for baseline LE8 score.

**Conclusions:** Sustained simultaneous adherence to high-quality diet and high PA through young adulthood was strongly and synergistically associated with high midlife CVH, supporting early, comprehensive, and sustained lifestyle prevention.

## Introduction

Cardiovascular disease (CVD) remains the primary global driver of premature mortality and chronic disability.^1^ Whereas clinical risk reduction strategies traditionally focus on midlife or older individuals, emerging longitudinal evidence underscores that young adulthood represents a critical life course window during which chronic pathophysiological processes develop and accelerate in the cardiovascular system. Habitual, structured moderate and/or vigorous intensity physical activity (or exercise) and optimal diet quality serve as foundational cornerstones in retarding subclinical cardiovascular deterioration via physiological pathways that help manage weight, maintain endothelial health, and improve blood pressure, lipid profiles, and blood sugar levels.^2^ Population data reveal that ideal levels of both diet and exercise remain strikingly low across most demographic groups in the United States.^3^ However, traditional epidemiological analyses frequently view these lifestyle components as static variables measured at a single point in time, failing to capture how behavioral habits shift, decline, or stabilize over decades.

Likewise, a major limitation in the current literature is the tendency to study dietary quality and physical activity as separate, non-interacting exposures. In a real-world setting, these lifestyle factors do not exist in a vacuum; they co-vary dynamically within an individual over time.^2^ By assessing these health behaviors independently, existing studies risk masking distinct behavioral phenotypes and missing potential synergistic biological effects, such as optimal nutrition amplifying the metabolic benefits induced by regular physical exertion.

Some investigators have mapped isolated health behaviors over extended periods^2^ but few have examined joint multi-trajectory patterns of evolution in diet and physical activity during early adulthood and their subsequent impact on mid-life CVH. Group-based dual trajectory modeling bridges this analytical gap by identifying latent, data-driven clusters of participants based on the parallel evolution of multiple behaviors over time.

To better quantify and track the multifaceted nature of cardiovascular health (CVH) in individuals and populations across the life course, in 2022 the American Heart Association introduced Life’s Essential 8^TM^ (LE8), a composite construct composed of diet, physical activity, nicotine exposure, sleep health, body mass index (BMI), blood lipids, blood glucose, and blood pressure.^4^ Higher LE8 scores have been linked consistently to greater longevity, compressed morbidity, markedly lower risk for all chronic diseases, and defense against systemic biological aging.^56^

The objective of this study was to investigate joint groups of dietary quality and physical activity through young adulthood and evaluate their associations with CVH across midlife. Using 35 years of prospectively collected data from the biracial Coronary Artery Risk Development in Young Adults (CARDIA) cohort, we sought to (1) stratify participants into distinct, data-driven latent classes based on joint evolution of Dietary Approaches to Stop Hypertension (DASH) scores and physical activity levels from the CARDIA Physical Activity History obtained at up to 7 exams from Exam Year 0 to Year 20 (mean age range at each exam 25 to 45 years), and (2) evaluate the prospective associations between these young adult behavioral trajectories and subsequent mid-life CVH patterns, as defined by LE8 scores, from Year 20 to Year 35 (mean age range at each exam 45 to 60 years).

## Methods

### Study Sample

The CARDIA study is a longitudinal, population-based cohort study begun in 1985-1986 with the enrollment of 5,115 Black or White (self-identified) men and women aged 18 to 30 years at 4 centers across the U.S. in Birmingham, AL, Chicago, IL, Minneapolis, MN, and Oakland, CA. CARDIA participants have undergone in-person examinations at baseline (Year 0: Y0) and at Y2, Y5, Y7, Y10, Y15, Y20, Y25, Y30, and Y35. Retention rates among surviving participants at each in-person examination were 91%, 86%, 81%, 79%, 74%, 72%, 72%, 71%, and 71% (during the COVID-19 pandemic), respectively. In addition to follow-up exams, contact is maintained with participants via telephone, mail, or email every 6 months, with annual interim medical history ascertainment. As of 2026, ∼85% of the surviving cohort members have been directly contacted and follow up for vital status is virtually complete through related contacts and intermittent National Death Index searches. Details of the study design and conduct have been published previously.^7^ Participants provided written informed consent and all study procedures have been approved by institutional review boards at the participating institutions.

### Diet and Physical Activity Ascertainment

Dietary intake over the past month was assessed with an interviewer-administered, quantitative diet history questionnaire data at exam years (Y) 0, 7 and 20. DASH (Dietary Approaches to Stop Hypertension) dietary score was computed for men and women separately, as follows. The average intakes from eight dietary groups of fruits, vegetables, whole grains, diary, nuts and legumes, red and processed meats, sodium, sweetened beverages were calculated for each participant. A quintile score (low=1 to high=5) for each mean intake of fruits, vegetables, whole grains, dairy, nuts and legumes was assigned to each participant and a quintile score (high=1 to low=5) was assigned for red and processed meats, sodium, sweetened beverages. The 8 quintile scores were summed to create a total score ranging from 8 to 40, where 8 indicated the lowest adherence and 40 indicated highest adherence to the DASH diet. Diet data from year 0 was carried forward to determine diet status for years 2 and 5, while year 7 diet data carried forward for years 10 and 15. Year 20 diet data were also carried forward to determine diet status for year 25 and afterward.

Physical activity (PA) was measured using the CARDIA Physical Activity History (PAH) recall questionnaire at every examination, which estimates moderate and vigorous intensity physical activity over the past 12 months via recall of 13 pre-specified activity types or categories within the leisure (n=12) or occupational (n=1) domains. For each activity type or category performed for at least one-hour during a month, participants reported the number of months (in the last 12 months). Of those, participants classified the reported number of months as “frequent”, which was operationalized individually for each activity type or category (ranges from 2-5 hours) or “less frequent”. The PAH is scored in exercise units (EU), which represents a weighted sum, based on the intensity level assigned to each activity type or category (ranges from 3-8 metabolic equivalents of task [METs]) and number of months of “less frequent” participation plus three-times the number of months of “frequent” participation. A prior study found that 300 EU is roughly equivalent to meeting U.S. recommendations for physical activity (i.e., 150 minutes per week of moderate or vigorous intensity PA).^8^ Diet quality and PA tend to co-vary, but they may differ within individuals. Group-based dual trajectory modeling was applied to identify latent classes of individuals with similar behaviors across diet and PA from Y0 to Y20. The exposure variable was dual trajectory grouping of PA combined with diet identified by a group-based multi-trajectory model with data collected from Y0 to Y20 (mean age ranges at each exam 25 to 45 years). Accordingly, participants were stratified into groups using multiple latent class analysis based on similar joint DASH and PA exposures from Y0 to Y20, as described below.

### CVH score

We quantified CVH according to the criteria established by the American Heart Association, including 4 health behaviors (diet, physical activity, smoking, and sleep) and 4 health factors (body mass index, non–high-density lipoprotein cholesterol level, blood glucose level, and blood pressure). Each component metric was assigned a score from 0 to 100 (higher indicates better CVH), and the overall LE8 score (range, 0 to 100 points) was calculated as the unweighted average of all available metrics.^4^ In primary analyses, LE8 scores were calculated using five components - nicotine exposure, body mass index, blood lipids, blood glucose, and blood pressure - excluding sleep health, diet, and physical activity due to data availability and to avoid mathematical circularity. As a sensitivity analysis, LE8 scores were recalculated using seven components (excluding only sleep health). We stratified participants into categorical groups by CVH status, with LE8 score ≥80 representing high CVH, 50–79 representing moderate CVH, and <50 representing low CVH.^4^ The primary outcome was LE8 trajectory groups identified by a group-based trajectory model from exam year 20, 25, 30, and 35 (mean age range at each exam 45 to 60 years).

### Exposure Covariates

Demographic variables including age, sex, race, and highest educational attainment, measured in years, were self-reported for all CARDIA participants. These covariates were adjusted for in all analyses.

### Statistical Analysis

For this study, participants were excluded if they had no LE8 trajectory outcome data (n = 2150), incomplete PA or diet data at baseline (n = 134), or incalculable PA and diet dual trajectory exposure data (n = 22). A minimum of three time points is required for latent-class trajectory analysis, so participants with fewer than three CVH assessments between Y20 and Y35 or fewer than three physical activity or diet assessments between Y0 and Y20 were excluded. In accordance with STROBE reporting guidelines, a total of 2,806 participants were included. Baseline characteristics of included and excluded participants are provided in Supplemental Table 1. Compared with included participants, those who were excluded tended to be slightly younger, and were more likely to be male and Black, have fewer years of education, and be smokers at baseline; there were minimal differences in other health behaviors and clinical measures.

Demographic and health characteristics were compared using analysis of variance for continuous and chi-square for categorical variables across dual trajectory groups of diet and PA. To examine the joint influence of diet quality and physical activity on cardiovascular health, we constructed a combined exposure variable representing concurrent adherence to the DASH diet and physical activity levels across exam cycles from Y0 to Y20. To characterize the dynamic patterns of these behaviors over time, we used multiple latent class modeling and estimated dual-trajectory models, which simultaneously model diet and physical activity trajectories and quantify the degree to which changes in one behavior are associated with changes in the other.^9^

We used the SAS procedure Proc Traj^10,11^ which applied latent class modeling to identify subgroups of participants with similar LE8 trajectories. Two trajectory groups were initially constructed and then expanded to three, four, and five trajectory groups. For each number of trajectory groups, polynomial orders for each trajectory shape were tested to identify the optimal fitting model. We chose the optimal number of dual trajectories based on the following criteria (1) >5% sample size in each multi-trajectory group (2) lowest Bayesian information criterion; (3) Average posterior probability values > 0.7 in each group to assign membership, as in prior analyses. A similar approach was applied in selecting optimal dual-trajectory latent classes for diet and PA. Age-, sex-, race- and education-adjusted multinomial logistic regression was used to model the association between diet-PA groups from Y0 to Y20 and CVH/LE8 trajectory patterns from Y20 to Y35.

## Results

### Sample Characteristics

A total of 2,806 participants were included in this secondary data analysis, of whom 1211 (43.2%) self-identified as male and 1,217 (43.4%) as Black. The mean (SD) baseline DASH diet score was 24.4 (4.9) and mean (SD) baseline physical activity intensity score was 420.4 (293.5).

Four joint diet/PA trajectory exposure groups across Y0 to Y20 were identified **(Figure 1)** using the multiple latent class approach and they were labeled subjectively as: Group 1, low DASH/very low PA (10.9%); Group 2, low DASH/moderate-declining PA (25.6%); Group 3, moderate DASH/high-declining PA (41.5%); Group 4, high DASH/high-declining PA (22.0%). **Table 1** shows descriptive characteristics of study participants overall and stratified by joint diet and PA groups. Diet-PA Group 1 was comprised predominantly of Black women while Diet-PA Group 4 was comprised largely of White women. The difference between the mean Group 1 and Group 4 DASH scores at baseline was 9.0 points and difference between the mean Group 1 and Group 4 PA scores at baseline was 346.6 units. Across both metrics, baseline LE8 scores demonstrated a modest gradient across groups from Group 1 to Group 4; however, the 5-component score yielded systemically higher values (ranging from 82.4 in Group 1 to 87.6 in Group 4) compared to the 7-component score (65.6 to 75.4, Table 1).

**Figure 1.**
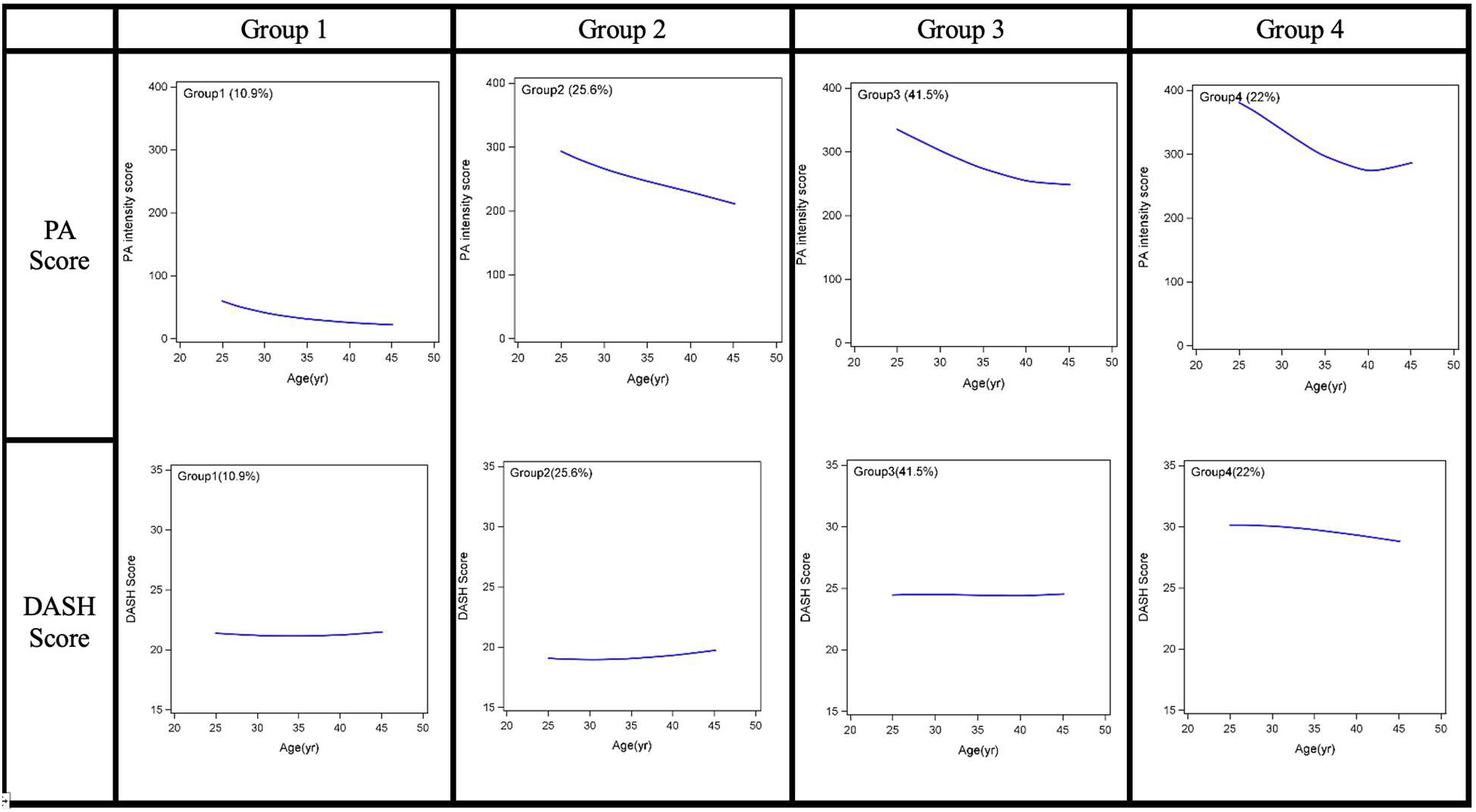
Joint groups of DASH score and PA intensity through young adulthood (Y0 to Y20, mean ages 25 to 45 years). Four trajectory groups were identified: low PA/moderate to low DASH (10.9%), moderate PA/low DASH (25.6%), moderate PA/moderate DASH (41.5%) and moderate PA/high DASH (22%).

**Table 1.** Characteristics of CARDIA Participants at Baseline (Year 0) Examination, Stratified by Joint DASH-PA groups.

|  | **Overall**<br>**(n=2806)** | **Group 1**<br>***Low DASH/***<br>***Very Low PA***<br>**(n=263)** | **Group 2**<br>***Low***<br>***DASH/Moderate***<br>***- declining PA***<br>**(n=658)** | **Group 3**<br>***Moderate***<br>***DASH/High-***<br>***declining PA***<br>**(n=1177)** | **Group 4**<br>***High***<br>***DASH/High-***<br>***declining PA***<br>**(n=708)** |
| --- | --- | --- | --- | --- | --- |
| Age, yrs, mean (SD) | 25.2 (3.6) | 24.3 (3.9) | 24.7 (3.6) | 25.1 (3.6) | 26.0 (3.1) |
| Male, N (%) | 1211<br>(43.2%) | 34 (12.9%) | 435 (66.1%) | 540 (45.9%) | 202 (28.5%) |
| Black, N (%) | 1217<br>(43.4%) | 212 (80.6%) | 429 (65.2%) | 470 (39.9%) | 106 (15.0%) |
| Total education, yrs,<br>mean (SD) | 16.0 (2.6) | 14.9 (2.3) | 15.0 (2.5) | 16.2 (2.5) | 17.1 (2.2) |
| Baseline DASH score,<br>mean (SD) | 24.4 (4.9) | 21.3 (3.1) | 19.2 (2.7) | 24.4 (2.8) | 30.3 (2.9) |
| Baseline PA score, mean<br>(SD) | 420.4<br>(293.5) | 142.1 (139.0) | 416.1 (298.7) | 443.9 (285.7) | 488.7 (285.3) |
| BMI, kg/m<sup>2</sup>, mean (SD) | 24.3 (4.6) | 25.3 (5.7) | 24.7 (4.8) | 24.4 (4.5) | 23.2 (4.1) |
| Total cholesterol, mg/dL,<br>mean (SD) | 177.4 (33.0) | 179.0 (35.4) | 179.0 (34.9) | 178.2 (32.8) | 174.1 (30.4) |
| HDL cholesterol, mg/dL,<br>mean (SD) | 53.6 (12.7) | 53.1 (12.0) | 52.5 (13.0) | 53.2 (12.5) | 55.4 (12.9) |
| Fasting glucose, mg/dL,<br>mean (SD) | 81.7 (8.9) | 81.3 (10.1) | 82.1 (9.9) | 81.4 (8.9) | 81.9 (7.4) |
| Systolic BP, mmHg, mean<br>(SD) | 109.7 (10.6) | 108.7 (9.7) | 112.2 (10.8) | 109.6 (10.8) | 108.0 (10.0) |
| Diastolic BP, mmHg,<br>mean (SD) | 68.5 (9.3) | 67.8 (9.0) | 69.4 (9.8) | 68.2 (9.5) | 68.2 (8.4) |
| BP medication use, N (%) | 47 (1.7%) | 8 (3.0%) | 13 (2.0%) | 20 (1.7%) | 6 (0.8%) |
| Current smoker, N (%) | 663 (23.7%) | 66 (25.5%) | 210 (32.0%) | 284 (24.2%) | 103 (14.6%) |
| LE8 at Y20-Y35 Traj |  |  |  |  |  |
| LE8 Score at Baseline Y0<br>(5 components) | 83.6 (12.4) | 82.4 (13.1) | 80.4 (12.8) | 83.1 (12.0) | 87.6 (11.5) |
| • High | 782 (27.9%) | 37 (14.1%) | 84 (12.8%) | 306 (26.0%) | 355 (50.1%) |
| • Moderate | 1354<br>(48.3%) | 135 (51.3%) | 354 (53.8%) | 590 (50.1%) | 275 (38.8%) |
| • Low | 670 (23.9%) | 91 (34.6%) | 220 (33.4%) | 281 (23.9%) | 78 (11.0%) |
| LE8 Score at Baseline Y0<br>(7 components) | 71.7 (10.0) | 65.6 (11.9) | 69.6 (10.3) | 72.0 (9.1) | 75.4 (8.7) |
| LE8 at Y20-Y35 Traj |  |  |  |  |  |
| • High | 762 (32.7%) | 10 (4.8%) | 97 (17.9%) | 329 (32.8%) | 326 (56.5%) |
| • Moderate | 1072<br>(46.0%) | 103 (49.3%) | 290 (53.6%) | 480 (47.8%) | 199 (34.5%) |
| • Low | 497 (21.3%) | 96 (45.9%) | 154 (28.5%) | 195 (19.4%) | 52 (9.0%) |

### Mid-life CVH trajectories

There were three distinct mid-life LE8 trajectories identified (low, moderate, and high CVH; **Figure 2**), whose LE8 scores differed at Y20 and did not overlap during follow up from Y20 to Y35. When looking at the CVH outcomes, 56% of individuals in group four achieved high cardiovascular health compared to just about 5% in group one. Conversely, about 46% in group one had low cardiovascular health compared to only about 9% of group 4 (Table 1).

**Figure 2.**
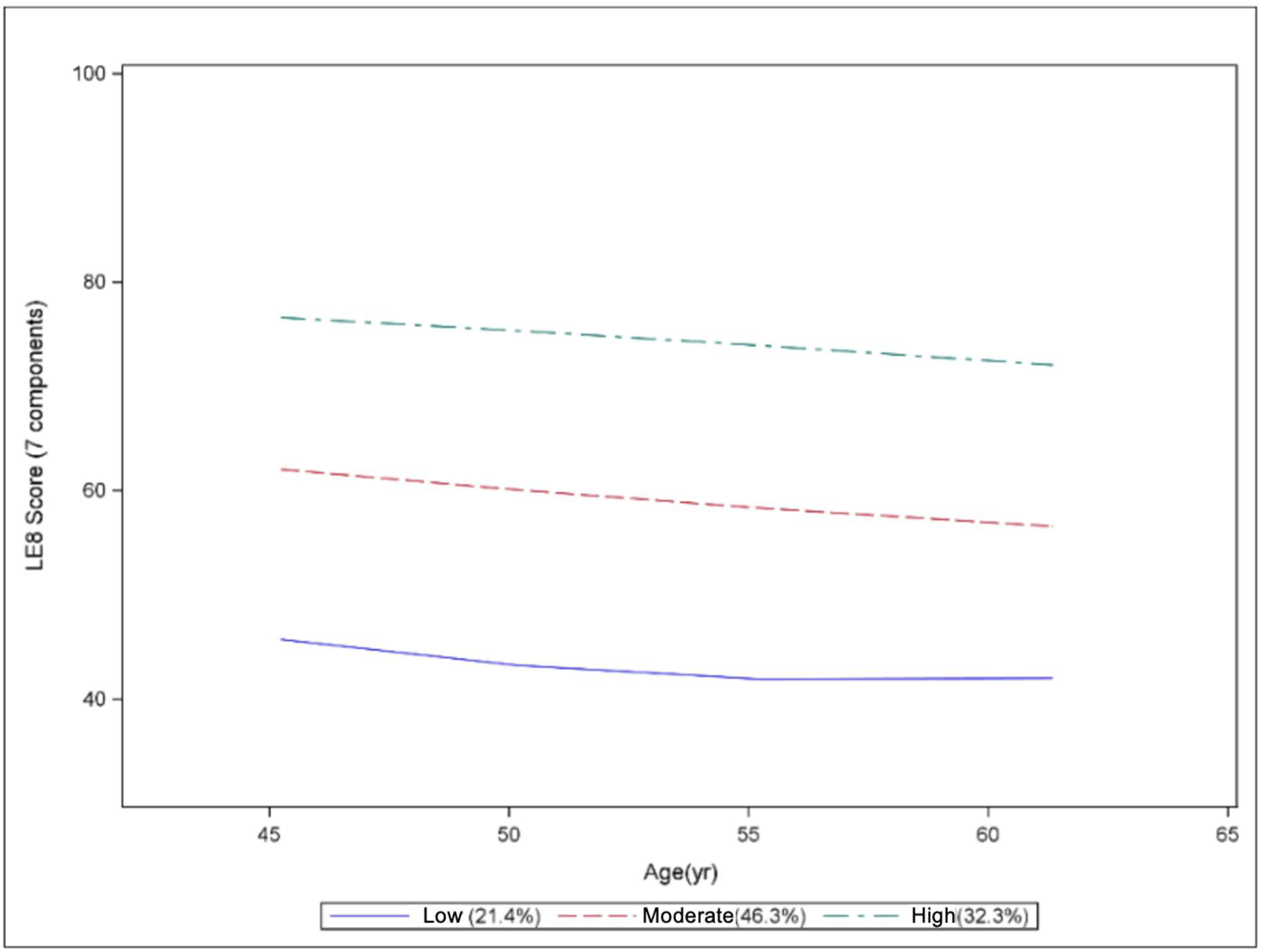
CVH LE8 Score Trajectories (Y20, 25, 30 and 35) From All LE8 Measures Excluding Sleep. Three patterns were identified: low, moderate and high CVH.

### Associations of Young Adult Diet-PA Group with Mid-Life CVH Status

Considering diet and PA jointly, participants in Groups 2, 3, and 4 had 4.8-, 8.6-, and 21.7-fold higher odds, respectively, of having high CVH compared to low CVH in midlife (**Table 2)**. Participants in Groups 2, 3, and 4 had 1.4-, 1.6-, and 2.2-fold higher odds, respectively, of having moderate CVH compared to low CVH in midlife.

**Table 2.** Age, sex, race and education status-adjusted odds ratios for mid-life CVH status (Y20 to Y35, mean ages 45 to 60) by joint DASH-PA Groups 1-4 during young adulthood (Y0 to Y20, mean ages 25 to 40) using 5 LE8 components.

|  | **Age, Sex, Race and Education Status Adjusted Odds Ratio for Membership in Mid-Life CVH Trajectory Group** |  |
| --- | --- | --- |
| **Young Adult Diet-PA Group** | **Moderate vs. Low CVH Trajectory** | **High vs. Low CVH Trajectory** |
| Group 1 | 1.0 (reference) | 1.0 (reference) |
| Group 2 | 1.36 (0.94, 1.96) | 4.83 (2.34, 9.98) |
| Group 3 | 1.58 (1.12, 2.23) | 8.61 (4.30, 17.2) |
| Group 4 | 2.15 (1.38, 3.35) | 21.7 (10.4, 45.6) |

These associations remained significant and similar after additional adjustment for the baseline (Y0) LE8 score and were consistent whether or not diet and PA metrics were incorporated in the midlife overall LE8 score trajectories (i.e. using 7 of the LE8 components in sensitivity analyses).

Individually, diet and PA at Y0 also had significant associations with midlife CVH (Supplemental Tables 2 and 3). Considering baseline DASH scores, participants in Quartiles 2, 3, and 4 of DASH score had 1.2-, 2.0-, and 3.1-fold higher odds, respectively, of having high CVH compared to low CVH in midlife. Across baseline PA scores, participants in Quartiles 2, 3, and 4 had 1.7-, 2.2-, and 2.9-fold higher odds, respectively, of having high CVH compared to low CVH in midlife.

## Discussion

In this study, we identified distinct joint trajectories of dietary quality and PA through young adulthood (mean age ranges by exam 20 to 45 years) that were strongly associated with midlife CVH status. Our approach represents a unique measure of capturing the simultaneous evolution of diet and physical activity over time, through joint trajectory modeling, rather than examining diet and physical activity as independent exposures or as single measures. This methodology allowed for identification of what participants tend to do in real life in reference to these two lifestyle factors together. Moreover, we were able to isolate specific subgroups, such as participants who adhered persistently to high physical activity but low dietary quality, that may have been otherwise obscured using traditional methods.

The primary finding of our analyses is the magnitude of association observed when diet and physical activity are modeled jointly rather than individually. Participants who maintained high diet quality and high physical activity (Group 4) simultaneously throughout young adulthood exhibited 21.7-fold higher odds of having high CVH in midlife, compared with those with consistently low diet quality and physical activity (Group 1). Notably, the association with high CVH appeared to be due to the combined effect of diet and physical activity. The odds ratio for joint high adherence for both diet and PA (21.74) was markedly higher than the odds ratio for high diet quality individually (7.2) or high physical activity individually (10.3). Given these findings were replicated with multiple dietary indexes to yield similarly significant results suggests that optimal nutrition, rather than a specific diet, and regular physical activity confer a level of physiological resilience that surpasses the benefits of a single lifestyle factor intervention.

Mechanistically, the synergistic association of diet and PA with high CVH in midlife has strong biologic plausibility. PA is known to enhance insulin sensitivity and endothelial function^15,16^ and high-quality diet provides the substrate availability and antioxidant capacity required to preserve endothelial integrity as well as metabolic homeostasis.^17^ Given the joint association was more than double that of either individual association, it seems plausible that there is a biologic interaction between dietary quality and physical activity such that high quality nutrition may potentiate the metabolic benefits of physical activity, and vice versa. This finding reemphasizes the conceptual CVH model of lifestyle factors being interconnected for optimal health. Future work should focus on identifying molecular mechanisms that may mediate this concurrent effect such as systemic inflammation and biological aging.

Furthermore, it is critical to note that achieving high CVH in midlife yields profound benefits that extend far beyond immediate disease prevention. Optimal midlife CVH is powerfully associated with greater overall longevity and a significantly extended healthspan.^5^ Rather than simply prolonging the later years of life, high CVH status acts to compress morbidity, effectively delaying the onset of age-related decline and concentrating the burden of illness into a much shorter window at the end of life.^12,13^ This systemic physiological resilience appears to serve as a robust defense mechanism against the broader biological processes of aging, resulting in markedly lower risks for all major chronic diseases of aging, including heart failure, cancer, type 2 diabetes, and dementia.^14^ Ultimately, the synergistic effect of high CVH ensures that added years of life are characterized by sustained physical and cognitive function rather than prolonged disability.^14^ Our findings delineate the important association of maintaining high diet quality and high levels of PA through young adulthood to substantially increase the likelihood of having high CVH in midlife, and in turn reaping the benefits of high CVH across the remaining lifespan.

Our analysis showed that baseline scores were similar across all trajectory groups and the significant difference in midlife CVH between Groups 1-4 was driven by the persistence of joint diet-PA behaviors over time. This finding is clinically important as it highlights that young adulthood represents a malleable window in which 1) one can increase their odds of high CVH and 2) the cumulative trajectory of combined lifestyle behaviors exerts a stronger influence on future CVH than baseline status.

This study, to our knowledge, is one of the first to examine the joint effect of diet and physical activity as a predictor of midlife CVH. The strengths and limitations of this study should be noted. Strengths include a large, biracial study population with 35 years of follow-up. The use of latent class modeling, specifically dual trajectory modeling allowed us to examine dynamic behaviors over time rather than single cross-sectional measures. The same repeated measures were used to estimate diet and physical activity exposure, allowing for consistency with the same measure, The primary analyses calculated LE8 scores using five of the LE8 physiologic metrics (nicotine exposure, body mass index, blood lipids, blood glucose, and blood pressure, in order to avoid including diet and PA as both exposures and outcomes initially) and were replicated with LE8 scores calculated using seven LE8 physiologic metrics (all excluding sleep). Limitations include measures of diet and physical activity being self-reported, which is susceptible to recall and reporting biases. Additionally, since sleep was not factored into either LE8 calculation, this reduces some comparability with full LE8 score studies. Given the observational nature of the study, causal inference may be limited; however, numerous interventional studies have established causal relationships for diet and PA with most of the other CVH metrics.

## Conclusions

Our findings demonstrate the substantial importance of diet and PA, particularly maintaining a combination of high-quality diet and greater PA, through young adulthood for mid-life CVH. Simultaneous adherence to healthy diet and high physical activity through young adulthood was associated with 21.7-fold higher odds of having high CVH in midlife, even after adjustment for baseline LE8 score, highlighting the importance of initiating lifestyle prevention strategies early that are comprehensive and sustained. Public health messaging and clinicians should focus on the synergistic value of combining dietary quality with physical activity for health promotion and cardiovascular disease prevention.

## Data Availability

The data analyzed in this study were obtained from the Coronary Artery Risk Development in Young Adults (CARDIA) Study, supported by the National Heart, Lung, and Blood Institute (NHLBI). De-identified individual-level participant data from the CARDIA study are available to qualified researchers through the NHLBI Biologic Specimen and Data Repository Information Coordinating Center (BioLINCC) or via application to the CARDIA Coordinating Center. Access to the dataset is subject to approval of a data distribution agreement and institutional review board requirements. The analytic code used for the group-based dual-trajectory modeling and statistical analyses in this study is available from the corresponding author upon reasonable request

## Acknowledgements

The Coronary Artery Risk Development in Young Adults Study (CARDIA) is conducted and supported by the National Heart, Lung, and Blood Institute (NHLBI) in collaboration with the University of Alabama at Birmingham (75N92023D00002 & 75N92023D00005), Northwestern University (75N92023D00004), University of Minnesota (75N92023D00006), and Kaiser Foundation Research Institute (75N92023D00003). This manuscript has been reviewed by CARDIA for scientific content.

## Supplemental Material

**Supplemental Table 1.** Characteristics of CARDIA Participants at Baseline (Year 0) Examination, Stratified by Inclusion/Exclusion in Current Analysis.

|  | **All (n = 5112)** | **Excluded**<br>**(n=2306)** | **Included**<br>**(n=2806)** |
| --- | --- | --- | --- |
| Age, yrs, mean (SD) | 24.8 (3.7) | 24.5 (3.7) | 25.2 (3.6) |
| Male, N (%) | 2327 (45.5%) | 1116 (48.4%) | 1211 (43.2%) |
| Black, N (%) | 2637 (51.6%) | 1420 (61.6%) | 1217 (43.4%) |
| Total education, yrs, mean (SD) | 15.3 (2.7) | 14.5 (2.6) | 16.0 (2.6) |
| Baseline DASH score, mean (SD) | 23.9 (4.9) | 23.4 (4.8) | 24.4 (4.9) |
| PA intensity score, mean (SD) | 420.1 (300.8) | 419.7 (309.5) | 420.4 (293.5) |
| BMI, kg/m<sup>2</sup>, mean (SD) | 24.5 (5.0) | 24.8 (5.5) | 24.3 (4.6) |
| Total cholesterol, mg/dL, mean (SD) | 176.8 (33.5) | 175.9 (34.0) | 177.4 (33.0) |
| HDL cholesterol, mg/dL, mean (SD) | 53.2 (13.2) | 52.7 (13.8) | 53.6 (12.7) |
| Fasting glucose, mg/dL, mean (SD) | 82.6 (16.3) | 83.7 (22.2) | 81.7 (8.9) |
| Systolic BP, mmHg, mean (SD) | 110.4 (11.0) | 111.3 (11.3) | 109.7 (10.6) |
| Diastolic BP, mmHg, mean (SD) | 68.6 (9.6) | 68.7 (10.0) | 68.5 (9.3) |
| BP medication use, N (%) | 115 (2.2%) | 68 (2.9%) | 47 (1.7%) |
| Current smoker, N (%) | 1544 (30.4%) | 881 (38.6%) | 663 (23.7%) |

**Supplemental Table 2.** Age, sex, race and education status-adjusted odds ratios for mid-life CVH status (Y20 to Y35, means ages 45 to 60) by Baseline PA Quartiles.

|  | Age, Sex, Race and Maximal Education Status Adjusted Odds Ratio for CVH Trajectory Status by Baseline Physical Activity Scores |  |
| --- | --- | --- |
|  | Moderate vs. Low CVH | High vs. Low CVH |
| PA Quartile 1 | 1.0 (reference) | 1.0 (reference) |
| PA Quartile 2 | 1.21 (0.901, 1.621) | 1.65 (1.158, 2.350) |
| PA Quartile 3 | 1.21 (0.886, 1.647) | 2.20 (1.533, 3.148) |
| PA Quartile 4 | 1.37 (0.978, 1.911) | 2.87 (1.958, 4.206) |
|  | Adjusted for Baseline LE8 Measure |  |
| PA Quartile 1 | 1.0 (reference) | 1.0 (reference) |
| PA Quartile 2 | 1.22 (0.883, 1.6) | 1.67 (1.124, 2.488) |
| PA Quartile 3 | 1.25 (0.894, 1.753) | 2.25 (1.498, 3.379) |
| PA Quartile 4 | 1.35 (0.937, 1.937) | 2.79 (1.816, 4.295) |

**Supplemental Table 3.** Age, sex, race and education status-adjusted odds ratios for mid-life CVH status (Y20 to Y35, means ages 45 to 60) by Baseline DASH Quartiles.

|  | Age, Sex, Race and Maximal Education Status Adjusted Odds Ratio for CVH Trajectory Status by Baseline DASH Dietary Scores |  |
| --- | --- | --- |
|  | Moderate vs. Low CVH | High vs. Low CVH |
| DASH Quartile 1 | 1.0 (reference) | 1.0 (reference) |
| DASH Quartile 2 | 0.95 (0.72, 1.26) | 1.18 (0.828, 1.680) |
| DASH Quartile 3 | 1.09 (0.79, 1.53) | 1.99 (1.35, 2.93) |
| DASH Quartile 4 | 1.21 (0.84, 1.74) | 3.13 (2.09, 4.67) |
|  | Adjusted for Baseline LE8 Measure |  |
| DASH Quartile 1 | 1.0 (reference) | 1.0 (reference) |
| DASH Quartile 2 | 1.08 (0.80, 1.46) | 1.41 (0.95, 2.09) |
| DASH Quartile 3 | 1.17 (0.817, 1.666) | 2.03 (1.32, 3.13) |
| DASH Quartile 4 | 1.10 (0.74, 1.62) | 2.63 (1.68, 4.10) |

## Notes

### Competing Interest Statement

The authors have declared no competing interest.

### Author Declarations

The CARDIA study protocol was approved by the Institutional Review Boards (IRBs) of all four field centers (University of Alabama at Birmingham, Northwestern University, University of Minnesota, and Kaiser Foundation Research Institute) and the CARDIA Coordinating Center, and written informed consent was obtained from all participants. Analysis of secondary, de-identified CARDIA data for the present study was approved by the CARDIA committee and IRB exempt.

